# Household Hardships and Responses to COVID-19 Pandemic-Related Shocks in Eastern Ethiopia

**DOI:** 10.1101/2023.02.01.23285322

**Authors:** Jonathan A. Muir, Merga Dheresa, Zachary J. Madewell, Tamirat Getachew, Gamachis Daraje, Gezahegn Mengesha, Cynthia G. Whitney, Nega Assefa, Solveig A. Cunningham

## Abstract

**Background:** COVID-19 resulted in enormous disruption to life around the world. To quell disease spread, governments implemented lockdowns that likely created hardships for households. To improve knowledge of consequences, we examine how the pandemic period was associated with household hardships and assess factors associated with these hardships.

**Methods:** We conducted a cross-sectional study using quasi-Poisson regression to examine factors associated with household hardships. Data were collected between August and September of 2021 from a random sample of 880 households living within a Health and Demographic Surveillance System (HDSS) located in the Harari Region and the District of Kersa, both in Eastern Ethiopia.

**Results:** Having a head of household with no education, residing in a rural area, larger household size, lower income and/or wealth, and community responses to COVID-19, including lockdowns and travel restrictions, were independently associated with experiencing household hardships.

**Conclusions:** Our results identify characteristics of groups at-risk for household hardships during the pandemic; these findings may inform efforts to mitigate the consequences of COVID-19 and future disease outbreaks.

## Introduction

The COVID-19 pandemic resulted in enormous disruptions to life around the world. Beyond direct health effects [1], the pandemic had social and economic consequences as government-enforced lockdowns were implemented to stem the pace and severity of the disease [2]. To adhere to these lockdowns, many businesses closed their in-person workplaces temporarily; for some, business closure was permanent. Mitigation efforts also affected healthcare providers, who had to restrict in-person access to patients and/or limit services. These efforts may have affected the health of young children, pregnant women, and mothers by limiting access to healthcare and food, especially in isolated regions of resource-limited countries [3–18]. These indirect effects, in return, would exacerbate direct effects of the SARS-CoV-2 virus by increasing risk for undernutrition and other conditions that heighten the danger of serious illness [6, 18]. Knowledge of the extent to which households living in resource-limited countries have experienced resource restrictions and other hardships remains limited due to incomplete or nonexistent population surveillance [19].

In sub-Saharan Africa (SSA), national governments undertook considerable efforts to quell the spread of COVID-19 [2, 20]. Mitigation efforts in Ethiopia included social distancing, lockdowns, and emphasizing hygiene protocols; these efforts began on March 16*^th^* of 2020, intensified on March 20*^th^*, and ultimately a five-month-long state of emergency was declared on April 10*^th^* [21]. Economic and social disparities across different sociodemographic groups and geospatial inequalities may have resulted in the uneven implementation of these efforts [8]. These disparities may also have resulted in differential household and community vulnerability to unintended consequences of these efforts.

Vulnerability is the collective effect of cultural, economic, institutional, political, and social processes that modify the experience of and recovery from a given hazard [22]. In the context of disasters, it is often not the hazard itself that creates the disaster; rather, the disaster is the impact on individual and community coping patterns and the inputs and outputs of social systems [23–25]. Social vulnerability is partially the result of social disparities that shape or influence the susceptibility of different groups to hazards while also controlling their capacity to respond [26, 27]. Individual and household level factors often associated with vulnerability include demographic characteristics such as age, ethnicity, race, and sex; socioeconomic status (e.g., lower income, wealth, employment, and/or education); household composition (e.g., presence of children or elderly); and housing and transportation [28]. Social vulnerability also involves place disparities stemming from characteristics of communities and the built environment. For example, differential availability of scarce resources between urban and rural areas may exacerbate individual- and household-level vulnerabilities to hazards [29–31]. To understand the broader consequences of the pandemic, it is important to consider economic, political, and social markers of vulnerability at the individual, household, and community level [23–27].

In Ethiopia, those already burdened by social and economic disparities and limited in their ability to access resources are at greater risk for experiencing additional hardships during the pandemic: older adults, people with disabilities or pre-existing medical conditions, the poor, people living in congested residences and/or slums, pregnant women, and the unemployed [8, 10, 20]. Rural populations in Ethiopia are at elevated risk for food insecurity and agricultural hardships as climate change, severe drought, conflict, and environmental degradation have culminated in societal shocks affecting livelihoods, particularly for farmers [32, 33]. Populations lacking access to safe drinking water and/or sanitary environments, reliant upon emergency food aid, or with limited access to media or other communication technologies are also at greater risk of adverse consequences of the pandemic [20]. Vulnerability requires context-specific interventions to address indirect costs of the pandemic [34]. We used data from an existing Health and Demographic Surveillance System (HDSS) in Eastern Ethiopia to analyze the prevalence of hardships experienced during the pandemic and examine factors associated with vulnerability to hardships. We explore household responses to these hardships and analyze factors associated with households using these coping strategies.

## Methods

### Study Setting

The setting for this study is a predominantly rural area in the Kersa District and an urban area in the Harari People’s National Regional State in Eastern Ethiopia [35, 36]. The rural area consists of 24 kebeles (a neighborhood or ward) and covers 353 km^2^, with a population of 135,754 in 25,653 households. The urban area consists of 12 kebeles, a population of 55,773 in 14,768 households, across 25.4 km^2^. The population has been followed through a Health and Demographic Surveillance System since 2012, with demographic and health-related information regularly collected (see Figure 1).

**Figure 1.**
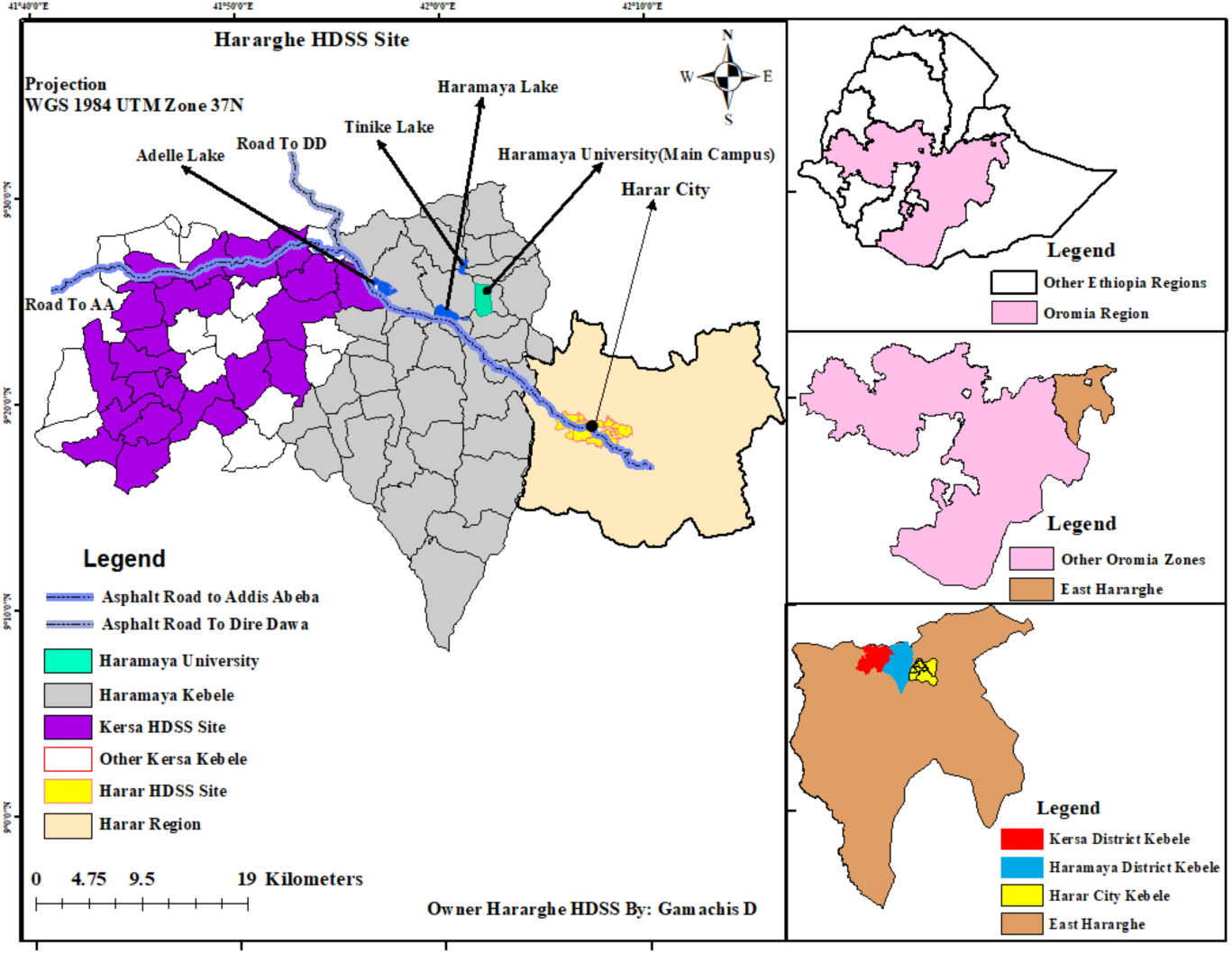
The Harar and Kersa Health and Demographic Surveillance System (HDSS) in Eastern Ethiopia. The smaller map panels on the right identify the location of the HDSS catchment areas within Eastern Ethiopia. Maps were created by the research team using shapefiles from the UN Humanitarian Data Exchange hosted by DataHub [37].

### Study Design

This study is part of a larger examination within the Child Health and Mortality Prevention Surveillance (CHAMPS) network to understand the consequences of COVID-19 lockdowns for child health and mortality [12–14]. We administered a short questionnaire designed to understand how the pandemic and related lockdowns may have affected the livelihood, food availability, and healthcare of households. Households were selected from a sample frame of all households in the HDSS using simple random sampling to achieve a sample size of 440 from the rural and from the urban catchment areas (total sample size of 880). The sample size was specified to detect prevalence of changes in accessing healthcare. A priori specifications were 50% of the population experiencing changes, 95% CI, precision of 0.05 and non-response adjustment of 10%.

The questionnaire was structured into five sections: knowledge regarding the spread of COVID-19; food availability; COVID-19-related shocks and coping; underfive child healthcare services; and healthcare services for pregnant women. Questions in the survey instrument related to hardships associated with the pandemic period asked respondents to consider whether a given hardship had occurred since March 2020. Data collectors were drawn from the fieldwork teams of HDSS enumerators already trained and working in the HDSS. Data collection occurred between August and September 2021 and was carried out through tablet-based in-person interviews with adult household members. All of the 880 sampled households consented and participated in the survey. Data from the questionnaire were linked with data from the most recently completed HDSS round (collected from January to May 2020) to incorporate additional demographic data about the sampled household, specifically: age, sex, occupation, and education of the head of household; the number of children under 5 years of age and the number of adults over age 60 in the household; and household assets and residence construction materials. Data quality assurance and cleaning followed standard procedures for the HDSS [35, 38]. Inconsistent or missing data were flagged for data collectors to correct. Field supervisors and the field coordinator selected a random sub-sample of questionnaires for re-visits to validate the recorded information. Implementation of the module was approved by the Institutional Health Research Ethics Review Committee (IHRERC); approval reference number Ref.No.IHRERC/127/2021. The data from the Ethiopia COVID- 19 lockdown questionnaire are publicly available through the CHAMPS Population Surveillance Dataverse and have been described in greater detail elsewhere [39, 40].

### Measures

The primary outcome variable, Household Hardships, was generated as an additive index of the number of hardships a household reported experiencing since the onset of the COVID-19 pandemic (i.e., during the 6 to 7-month period since March 2020); it was coded as a count variable ranging from 0 to 9, with 1 point given for each hardship a household reported from the list in Figure 2. Information on household hardships were gathered from the following survey question: “Has your household been affected by any of these events since mid-March?” Responses included: job loss; business closure; disruption of farming; disruption of livestock activities; disruption of fishing activities; increased price of farming or business inputs; decreased price of farming or business outputs, increased price of major food items consumed; and illness, injury, or death of any household member. The percentage of households that reported a given hardship is presented in Figure 2. The additive index had a Cronbach’s alpha score of 0.77 signifying high internal consistency [41].

**Figure 2.**
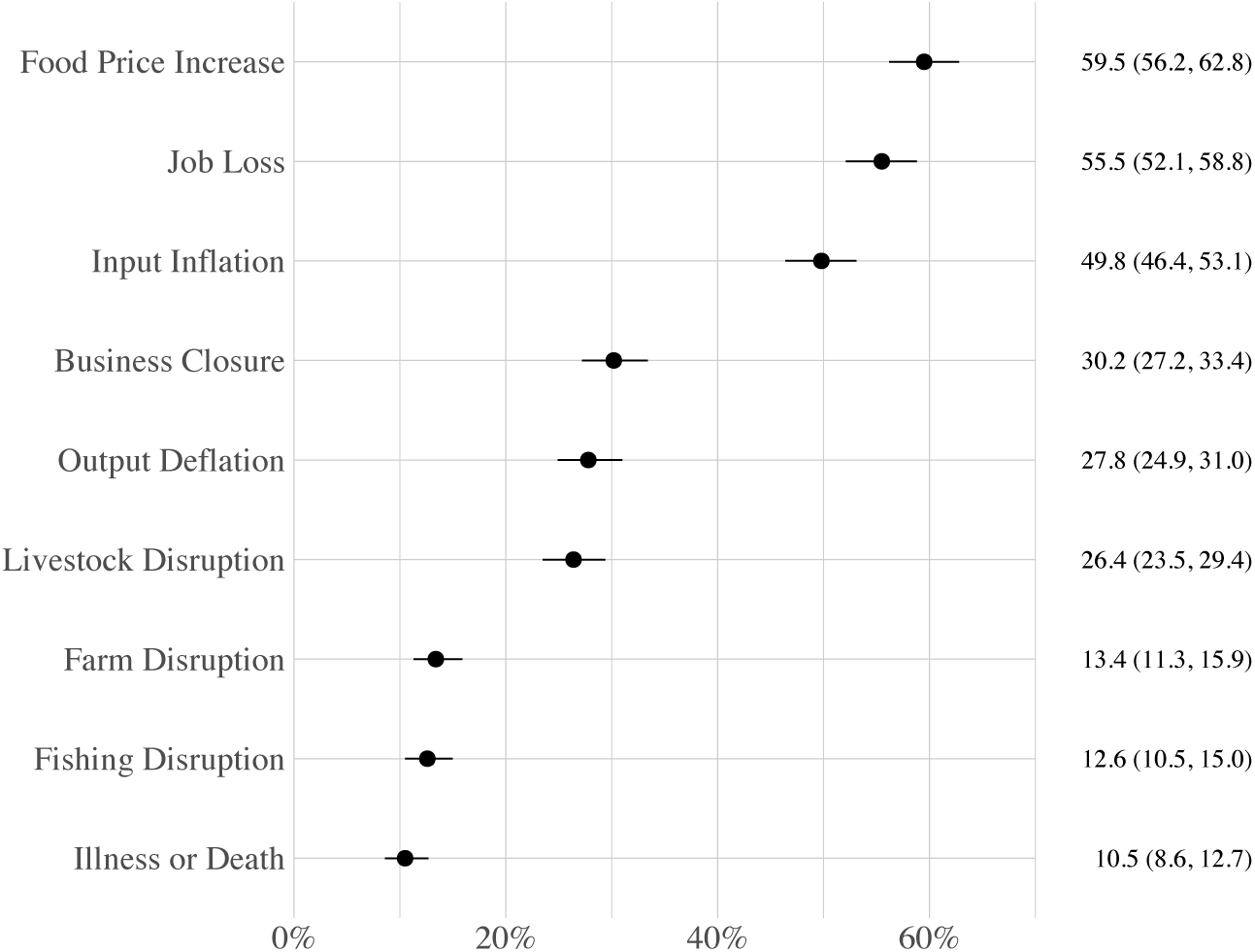
Percentage (95% confidence interval) of Households that Reported a Given Hardship. Represents the distribution of hardships used in the additive index Household Hardships (n = 880).

As an alternative representation of hardships used in a sensitivity analysis, we used principal components analysis (PCA) with varimax rotation to extract orthogonal factors from the variables in Household Hardships. The eigenvalues of the correlation matrix from PCA demonstrated that the first factor explained 40% of the variability in the data and the second explained 20%. Subsequent factors explained little variability. We opted to retain the first factor, which included job loss, business closure, farm disruption, livestock disruption, fishing disruption, and output deflation, as an index we termed Household Disruptions. Variables highly correlated with the factor were weighted against their eigenvector coefficients. Details related to the PCA results for the Household Disruptions are presented in Appendix A (see Table A.1).

Additional outcome variables were analyzed in a secondary analysis for households that reported experiencing at least one hardship during the pandemic. These outcome variables represented a variety of strategies that households may have employed in an attempt to mitigate experiences of hardship during the pandemic (e.g., household reduction of food consumption or the selling of assets); they were all coded Yes = 1 and No = 0 to signify if a given strategy was employed. A complete list of these additional outcome variables and corresponding descriptive statistics are presented in Figure 7.

Right-hand side variables in our analyses included information on the head of household, characteristics of the household and its homestead, and information related to household awareness of local government responses to COVID-19. Head of household variables included age (coded as 1 = 15 to 40 years, 2 = 41 to 50 years, 3 = 51 to 60 years, and 4 = *>*60 years); sex (coded as male = 0 and female = 1); ethnicity (coded as Amhara = 1 (reference) and other = 2, which included Oromo, Somali, Gurage, Harari and Tigray); education (coded as no formal education = 0 (reference), any level of education = 1); occupation (coded as 1 = farmer/domestic (reference), 2 = student, 3 = professional, 4 = sales, 5 = daily laborer, 6 = other employment, 7 = unemployed/retired). Household variables were urbanicity (coded as urban (Harar) = 1 and rural (Kersa) = 0); household size (coded as 1 = 1-2 individuals (reference), 2 = 3-4 individuals, 3 = 5-6 individuals, 4 = 7-8 individuals, 5 = 9+ individuals); children under age 5 (coded as 0 = no, 1 = yes); adults over age 60 (coded as 0 = no, 1 = yes); monthly income (coded as 1 = 0-1,200 Birr Ethiopia Birr (reference), 2 = 1,201-2,000 Birr, 3 = 2,001-3,000 Birr, 4 = 3,001-4,600 Birr, and 5 = *>*4,600 Birr; based on exchange rates at the time of the survey, this is roughly equivalent to 1 = Less than 33 USD (reference), 2 = 33 to 55 USD, 3 = 55 to 83 USD, 4 = 83 to 127 USD, and 5 = More than 127 USD (overlap in USD ranges due to exchange rate resulting in small differences in USD); and a wealth index based on a list of household assets (coded as an 1 = poorest (reference), 2 = poorer, 3 = middle, 4 = richer, 5 = richest). See Appendix B for information on the principal components analysis used to generate the wealth index. Covid response variables were used to analyze associations between household hardships and a household’s subjective awareness of community/government interventions that were implemented to stem the spread of COVID-19. Responses were gathered from the following self-report survey question: “What steps has your community/government taken to curb the spread of the coronavirus in your area?” Intervention variables included lockdowns, travel restrictions, business closures, and intervention centers; each coded as Yes = 1 and No = 0.

### Analytic Strategy

Data cleaning and analysis was performed using R version 4.2.0 [42]. Education and occupation data from the HDSS were missing for nine household heads; these households were removed from analyses that included these variables. A series of unadjusted quasi-Poisson regression models were used to analyze associations between right-hand side variables and either the additive index *Household Hardships* or the PCA-based index of *Household Disruptions* (results from the unadjusted analyses are available upon request) [43]. Adjusted quasi-Poisson regression models were then used to control for the other characteristics. We assessed model fit by comparison of Akaike Information Criterion (AIC) and Bayesian Information Criterion (BIC) scores—both scores were the smallest for the fully adjusted model, indicating that this model had superior fit compared to simpler or pathway-specific models. Results are reported as Adjusted Incidence Rate Ratios (AIRR) with 95% confidence intervals (95% CI) and visualized with forest plots [44, 45]. Given anticipated differences in demographic, economic, and social characteristics between populations living in urban vs. rural areas, effect modification was evaluated using interaction terms and visualized as predicted counts. A secondary analysis was conducted for those households that reported experiencing at least one hardship during the pandemic. This analysis used adjusted logistic regression to examine factors associated with household responses to pandemic hardships. We present results for the response strategies that were used by at least 10% of households.

## Results

Only 13% of households reported not experiencing any of the measured hardships; the remaining 87% experienced at least one hardship (Figure 3). The average number of hardships experienced during the pandemic was 2.86, with a standard deviation of 2.31; the interquartile range spanned 1 to 4 hardships. A majority of households (59.5%) observed an increase in local food prices (see Figure 2). Other common hardships included a household member losing their job (55.5%), observing increases in the cost of inputs for businesses or farms (49.8%), business closures (30.2%), and reductions in the value of business or farming outputs (27.8%).

**Figure 3.**
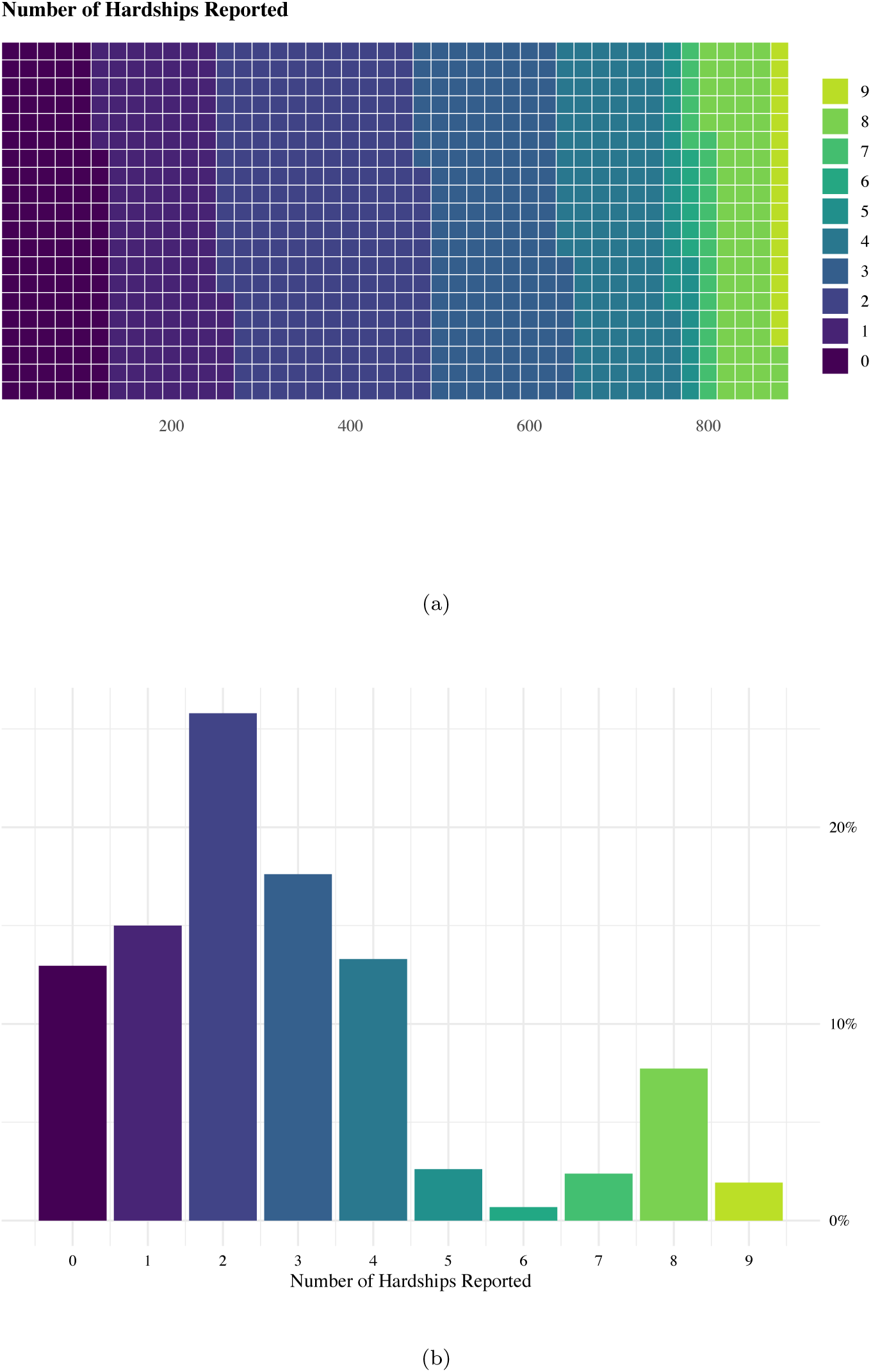
Number of Hardships Experienced during the COVID-19 Pandemic (n = 880). Figure 3a. presents a heat map of the number of hardships reported by the 880 sampled households. Figure 3b. presents the distribution of the additive index *Household Hardships*.

A majority of households (66%) were headed by a male household member (Table 1). Average age of the household head was 43 years, with a standard deviation of 15. Roughly a quarter of household heads were from the Amhara ethnic group. A majority (58%) of household heads had obtained some level of education; their most common occupations were farmer (46%) and professional (27%). The median number of household members was 5, with a standard deviation of 2.4. Roughly a third of households had at least one child under the age of 5 and 27% of households had at least one family member over the age of 60. The most commonly reported government/community responses to the pandemic were business closures (48%) followed by travel restrictions (46%) and the establishment of isolation centers (44%).

**Table 1:** Descriptive Statistics for Head of Household, Household, and Local COVID-19 Response Characteristics (n = 880)

|  | Total | Urban | Rural |  |  |
| --- | --- | --- | --- | --- | --- |
|  | f (%) | f (%) | f (%) | χ<sup>2</sup> | p-value |
| Head of Household Characteristics |  |  |  |  |  |
| Age |  |  |  |  |  |
| 15-40 | 218 (25) | 105 (24) | 113 (26) | 0.8 | 0.853 |
| 41-50 | 265 (30) | 136 (31) | 129 (29) |  |  |
| 51-60 | 179 (20) | 87 (20) | 92 (21) |  |  |
| >60 | 218 (25) | 112 (25) | 106 (24) |  |  |
| Sex |  |  |  |  |  |
| Female | 303 (34) | 192 (44) | 111 (25) | 33.0 | <0.001 |
| Male | 577 (66) | 248 (56) | 329 (75) |  |  |
| Ethnicity |  |  |  |  |  |
| Amhara | 222 (25) | 192 ( 8.3) | 30 (7) | 158.1 | <0.001 |
| Other | 658 (75) | 321 ( 1.9) | 410 (93) |  |  |
| Education |  |  |  |  |  |
| Some Education | 510 (58) | 78 (18) | 283 (65) | 201.0 | <0.001 |
| No Education | 361 (41) | 360 (82) | 150 (35) |  |  |
| Occupation |  |  |  |  |  |
| Farmer/Subsistence | 401 (46) | 89 (18) | 312 (72) | 255.2 | <0.001 |
| Student | 59 (7) | 34 (82) | 25 (6) |  |  |
| Professional/Sales | 236 (27) | 186 (82) | 50 (12) |  |  |
| Daily Laborer | 67 (8) | 53 (82) | 14 (3) |  |  |
| Other work | 21 (2) | 8 (82) | 13 (3) |  |  |
| Unemployed/Retired | 87 (10) | 68 (82) | 19 (4) |  |  |
| Household Characteristics |  |  |  |  |  |
| Household Size |  |  |  |  |  |
| 1-2 | 146 (17) | 94 (21) | 52 (12) | 99.0 | <0.001 |
| 3-4 | 257 (29) | 168 (38) | 89 (20) |  |  |
| 5-6 | 247 (28) | 122 (28) | 125 (28) |  |  |
| 7-8 | 171 (19) | 37 (8) | 134 (30) |  |  |
| 9+ | 59 (7) | 19 (4) | 40 (9) |  |  |
| Children Under Age 5 |  |  |  |  |  |
| No | 578 (66) | 322 (73) | 256 (58) | 22.0 | <0.001 |
| Yes | 302 (34) | 118 (37) | 184 (42) |  |  |
| Adults Over Age 60 |  |  |  |  |  |
| No | 644 (73) | 306 (70) | 338 (77) | 6.0 | 0.015 |
| Yes | 236 (27) | 134 (30) | 102 (23) |  |  |
| Income (Birr) |  |  |  |  |  |
| >4600 | 172 (20) | 125 (28) | 49 (11) | 55.1 | <0.001 |
| 3001-4600 | 132 (15) | 71 (16) | 63 (14) |  |  |
| 2001-3000 | 165 (19) | 75 (17) | 93 (21) |  |  |
| 1201-2000 | 209 (24) | 73 (17) | 137 (31) |  |  |
| 0-1200 | 192 (22) | 96 (22) | 97 (22) |  |  |
| Wealth Index |  |  |  |  |  |
| Richest | 180 (20) | 95 (22) | 85 (19) | 13.4 | 0.009 |
| Richer | 174 (20) | 87 (20) | 87 (20) |  |  |
| Middle | 174 (20) | 72 (16) | 102 (23) |  |  |
| Poorer | 176 (20) | 81 (18) | 95 (22) |  |  |
| Poorest | 176 (20) | 105 (24) | 71 (16) |  |  |
| COVID-19 Community Responses |  |  |  |  |  |
| Business Closures |  |  |  |  |  |
| No | 461 (52) | 228 (52) | 233 (53) | 0.1 | 0.736 |
| Yes | 419 (48) | 212 (48) | 207 (47) |  |  |
| Isolation Centers |  |  |  |  |  |
| No | 497 (56) | 253 (58) | 244 (55) | 0.4 | 0.541 |
| Yes | 383 (44) | 187 (43) | 196 (45) |  |  |

Table 1: Descriptive Statistics for Head of Household, Household, and Local COVID-19 Response Characteristics (n = 880) (Continued)
|  |  |  |  |  |  |
| --- | --- | --- | --- | --- | --- |
| Lockdowns |  |  |  |  |  |
| No | 502 (57) | 264 (60) | 238 (54) | 3.1 | 0.077 |
| Yes | 378 (43) | 176 (40) | 202 (46) |  |  |
| Travel Restrictions |  |  |  |  |  |
| No | 474 (54) | 227 (52) | 247 (56) | 1.8 | 0.176 |
| Yes | 406 (46) | 213 (48) | 193 (44) |  |  |

Data source: CHAMPS Ethiopia Lockdown Module data and CHAMPS Ethiopia Health and Demographic Surveillance data. Notes: Nine observations were missing for head of household's education and occupation. One observation was missing for whether a household had experienced food insecurity prior to the onset of COVID-19. The "other" category for ethnicity included: Oromo, Somali, Gurage, Harari, Tigray, and "other". The methods for calculating the wealth index are described in Appendix B. Chi-square statistics and p-values were based on Chi-square tests evaluating potential differences in descriptive statistics by urban vs. rural residence.

After adjusting for other variables (Figure 4), the number of hardships experienced by households during the pandemic were significantly higher if the household resided in a rural area compared to an urban area (AIRR = 1.34, 95% CI [1.20, 1.49]). Compared to households with 1 to 2 household members, larger households experienced a significantly higher number of hardships during the pandemic; for example, for households with 7-8 members, the AIRR for experiencing hardships during the pandemic was 1.42 (95% CI [1.24, 4.65]). In contrast, households experienced a significantly lower number of hardships during the pandemic if the household head had at least some education compared to none (AIRR = 0.87, 95% CI [0.79, 0.96]); was employed in a professional occupation (AIRR = 0.75, 95% CI [0.66, 0.84]) or as a day laborer (AIRR = 0.74, 95% CI [0.62, 0.88]) compared to agriculture; had a monthly household income greater than 1,200 Birr (for example, the AIRR for experiencing hardships during the pandemic for households with a monthly income greater than 4,600 Birr was 0.57 (95% CI [0.49, 0.66])); or had more assets compared to the poorest households. Households also experienced a significantly higher number of hardships during the pandemic if they also reported local community/government implementation of lockdowns (AIRR = 1.29, 95% CI [1.14, 1.46]), travel restrictions (AIRR = 1.38, 95% CI [1.23, 1.56]), and the establishment of isolation centers (AIRR = 1.17, 95% CI [1.03, 1.33]) compared to households that did not report these local community/government responses. The age, sex, and ethnicity of the household head, as well as the presence of children under the age of 5 or adults older than age 60 in the household, were not associated with household hardships after adjusting for other variables. Results from the sensitivity analysis of the Household Disruptions index (Figure 5) were generally consistent with the results from analyzing the Household Hardships index.

**Figure 4.**
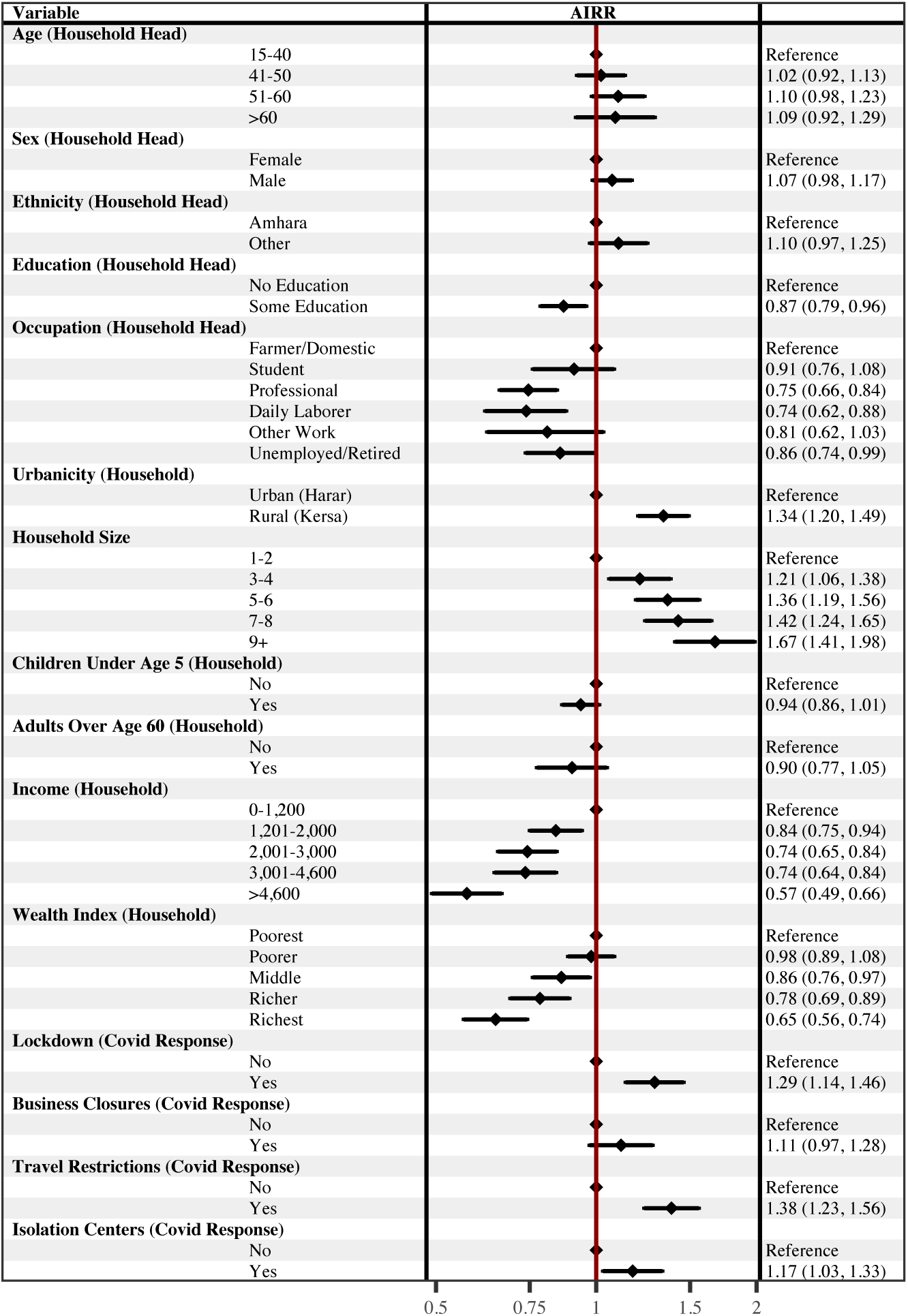
Adjusted Association with Household Hardships presented as Adjusted Incidence Rate Ratios (AIRR). Household Hardships was generated as an additive index of the number of hardships a household reported experiencing since the onset of the COVID-19 pandemic. The forest plot presents AIRRs with 95% confidence intervals from a multivariate quasi-Poisson regression model. The AIRRs were adjusted for the other variables included in the model. Education and Occupation had 9 missing values; Income had one outlier set to missing. (n=870)

**Figure 5.**
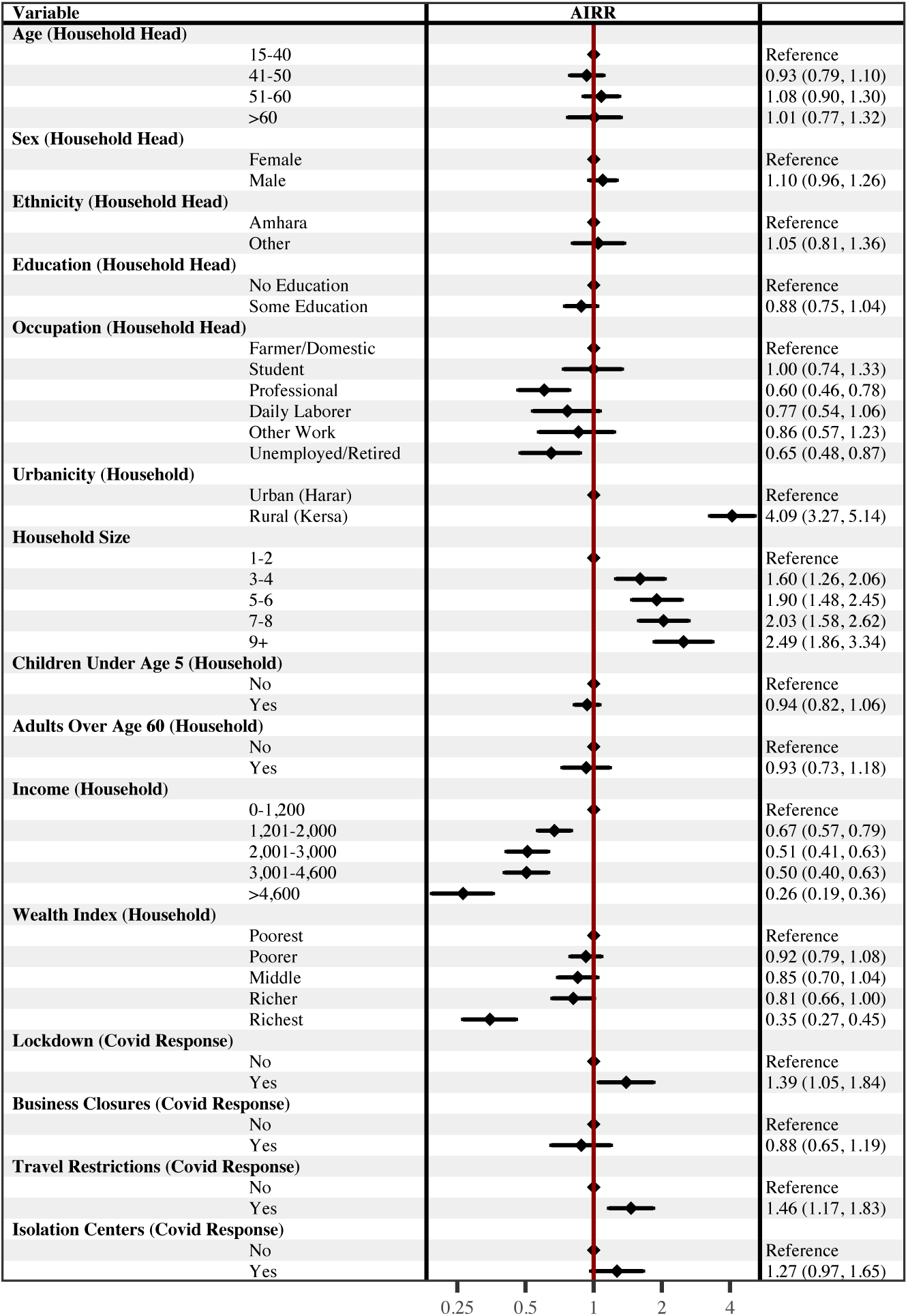
Adjusted Associations with Household Disruptions presented as Adjusted Incidence Rate Ratios (AIRR). Household Disruptions was generated as a PCA-based index of hardships a household reported experiencing since the onset of the COVID-19 pandemic. The forest plot presents AIRRs with 95% confidence intervals from a multivariate quasi-Poisson regression model. The AIRRs were adjusted for the other variables included in the model. Education and Occupation had 9 missing values; Income had one outlier set to missing. (n=870)

Baseline predicted counts of household hardships experienced during the pandemic based on urbanicity are presented as a facet grid in Figure 6, which also presents the predicted number of hardships estimated from models that included interaction terms between urbanicity and other variables. The top facet presents the elevated number of hardships experienced during the pandemic for households living in a rural compared to an urban area while holding all other variables at their mean. These baseline counts were estimated using results from the analytic model presented in Figure 5. The remaining facets present predicted counts estimated from models that included statistically significant interaction terms between urbanicity and other variables associated with Household Hardships while holding all adjusting variables at their mean. The predicted counts indicate that households headed by an individual who identified as an ethnic group other than the Amhara or who had no education were primarily at greater risk of experiencing hardships if they resided in a rural area than an urban area. Dose responses of a higher number of hardships experienced are seen for larger households, households with lower income, and households with lower wealth that were also residing in a rural area compared to an urban area. In contrast, while the overall number of hardships experienced was higher for households living in rural areas, local implementations of lockdowns and business closures as a response to the pandemic were only associated with a higher number of predicted household hardships for those households living in urban areas.

**Figure 6.**
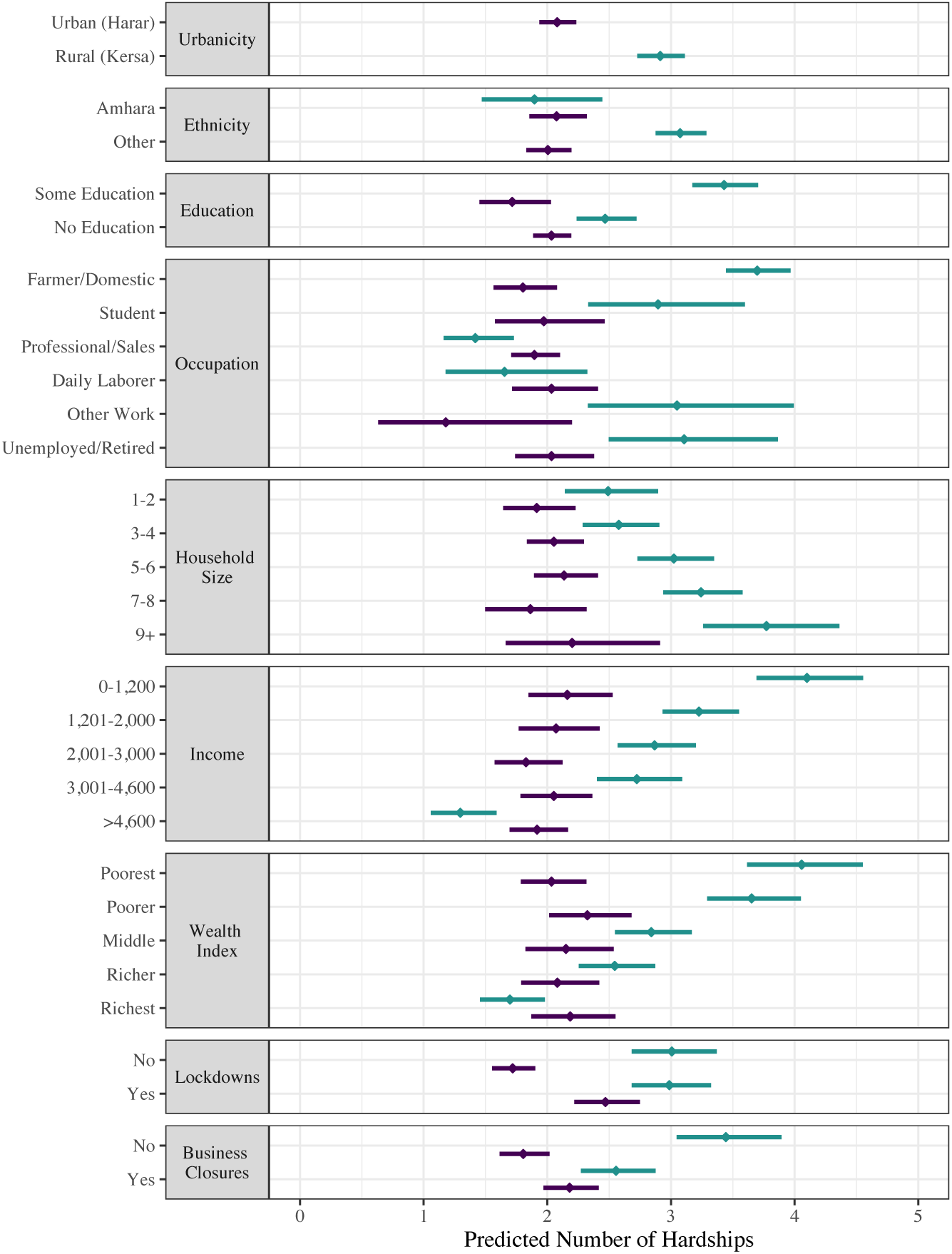
Evaluating Effect Modification between Urbanicity and Other Factors Associated with Household Hardships. The figure presents a facet grid of the predicted number of hardships households experienced from multiple analytic models. The top facet presents the predicted number of hardships experienced for households that resided in an urban (represented in green) vs. rural (represented in yellow) area, holding all other variables at their mean, to establish the baseline hardship count for households living in either of the two communities. The counts for this baseline were estimated using the results from the model presented in Figure 5. The remaining facets present the predicted number of hardships that households experienced based on estimates from models that included interaction terms between urbanicity, and other variables associated with Household Hardships while holding all adjusting variables at their mean.

Among households that reported experiencing at least one hardship during the pandemic, the most common household response was reducing food consumption, which was reported by 24.4% of affected households (see Figure 7). Other common responses included selling household assets (21.4%), seeking means to generate additional income (20.5%), seeking help from family or friends (14.0%), and borrowing from family or friends (10.6%). The median number of responses to hardships reported by households was one, with an interquartile range of 2 and a maximum number of 8 responses to hardships. However, a little more than half of all households (53%) did not report utilizing any of the measured responses.

**Figure 7.**
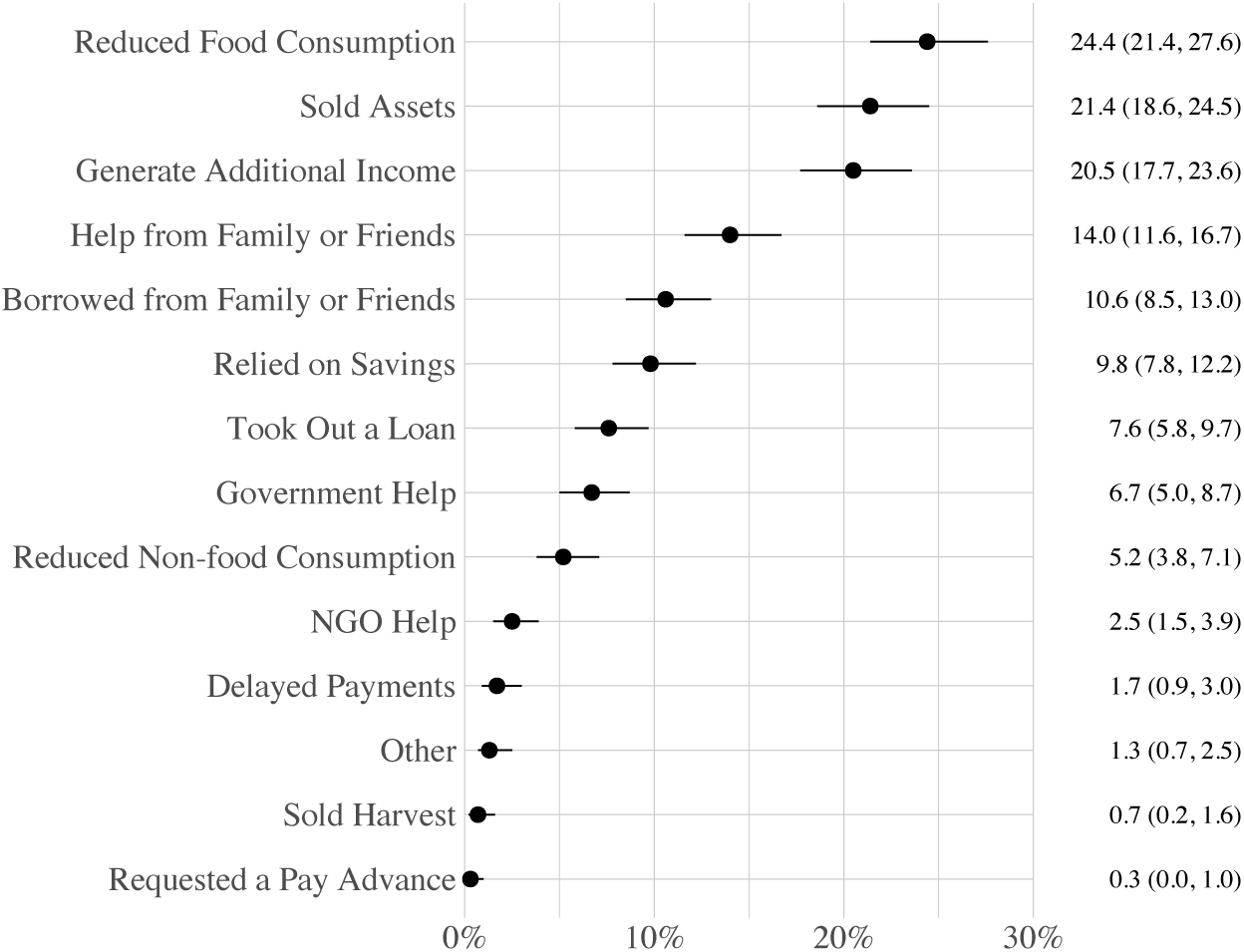
Percentage of Households that Utilized a Given Response to Hardships during the COVID-19 Pandemic (n = 766). Based on households that reported experiencing at least one hardship during the pandemic.

Detailed results are presented in Figures C.1 - C.5 in Appendix C. After adjusting for other variables, including other potential hardships, households that had a member lose their job were more likely to report reducing food consumption (AOR = 1.75, 95% CI [1.06, 2.92]); selling assets (AOR = 3.19, 95% CI [1.30, 8.41]); seeking to generate additional income (AOR = 2.82, 95% CI [1.55, 5.25]); seeking help from family and friends (AOR = 2.20, 95% CI [1.28, 3.83]); and borrowing from family or friends (AOR = 5.80, 95% CI [2.61, 13.82]). Households that observed increases in the costs of business or farming inputs were more likely to report reducing food consumption (AOR = 2.77, 95% CI [1.51, 5.20]); selling assets (AOR = 9.24, 95% CI [3.82, 23.96]); seeking help from family and friends (AOR = 3.45, 95% CI [1.80, 6.82]); and borrowing from family or friends (AOR = 5.54, 95% CI [2.30, 14.06]). Observing increased food prices was associated with households selling assets (AOR = 2.36, 95% CI [1.12, 4.96]). Households that had a member close their business during the pandemic were less likely to report selling assets (AOR = 0.08, 95% CI [0.02, 0.29]). Households that experienced farming disruptions were more likely to report seeking to generate additional income (AOR = 24.79, 95% CI [7.26, 97.71]). Households residing in a rural vs. an urban area were more likely to use all of the measured strategies to mitigate experiencing hardships associated with the pandemic; these associations were robust to adjusting for other variables.

## Discussion

In this study, we examined the prevalence of household hardships during the COVID-19 pandemic in a rural and an urban community in Eastern Ethiopia. We further examined demographic, economic, and social characteristics associated with these hardships as well as strategies households used to mitigate consequences of these hardships. Almost 90% of households reported experiencing at least one hardship since the onset of the pandemic; 75% of households reported experiencing at least 4 hardships. Risk factors for experiencing these hardships included demographic, economic, geographic, and social characteristics. Residing in a rural area magnified the strength of the associations for several household risk factors.

Households were more likely to report a higher number of hardships if the head of household had no education or worked as a farmer or in subsistence activities (housewife). Households were also more likely to report experiencing a higher number of hardships if they resided in a rural vs. an urban area, had a larger number of household members, had less monthly income, and were poorer compared to other households in the community. The pattern of association for both monthly income and household wealth is suggestive of a dose-response; households with less wealth and less monthly income were at significantly higher risk for experiencing a higher number of hardships. These associations are consistent with the vulnerability framework that we outlined, which identifies characteristics of individuals and households anticipated to increase the risk of negative outcomes in disaster situations [23–27]. The association between having a head of household employed in agricultural activities and households experiencing a higher number of hardships that include increased food insecurity may seem counterintuitive but is consistent with local conditions wherein climate change coupled with severe drought, conflict, and environmental degradation have culminated in societal shocks affecting rural livelihoods, particularly for farmers [32, 33].

Our findings are also consistent with risk factors commonly reported in studies on negative consequences of COVID-19 from other resource-limited countries, particularly in Africa, and other research from Ethiopia [5, 7, 8, 11, 12, 15, 20]. For example, a study from Kenya found that loss of employment, reductions in income, and food price increases were all associated with household hardships such as increased food insecurity [15]. Similarly, an early assessment of economic risks anticipated for households living in Ethiopia found that households with fewer assets, limited off-farm activities, and that lacked trading business were at heightened risk for significant welfare loss due to a slowdown in economic activities during the pandemic [20]. A study from Ethiopia found that the educational level of the household head, family size, and monthly income of the household were major determinants of rural households experiencing hardships such as food security [8]. Given the consistent pattern of these findings, interventions aimed at reducing the harmful effects of lockdowns and similar efforts should anticipate these characteristics as important risk factors and supplement aid to vulnerable groups.

Mitigation efforts in Ethiopia included social distancing, lockdowns, and emphasizing hygiene protocols [21]. We anticipated that geospatial inequalities may have resulted in differential household vulnerability to unintended consequences stemming from these interventions [8]. Our results are consistent with this premise and indicate that local community or government responses to the pandemic, such as lockdowns, travel restrictions, and the establishment of isolation centers, were associated with households experiencing a higher number of hardships. However, these associations varied to a degree by whether households resided in urban vs. rural areas; households living in urban areas reported experiencing a higher number of hardships if they also reported local implementation of these intervention efforts. These observations may stem from having humanitarian assistance policies and programs established in rural areas of Ethiopia that increase the resilience of rural households, but corresponding policies and programs for urban areas are lacking [46, 47]. The importance of having established, functioning social safety nets was observed in a Kenya study that found a heightened vulnerability amount urban households [15]. Interventions to reduce the harmful effects of lockdowns and similar efforts to limit disease spread should anticipate the need for increased vulnerability among urban populations.

Households that experienced hardships during the pandemic relied upon a variety of coping strategies to mitigate harmful effects; implementation of these strategies varied to a degree by the type of hardship a household experienced. Households that experienced the loss of employment by a household member or observed increasing costs of business or farming inputs were more likely to utilize the broadest combination of household responses to pandemic hardships. Compared to urban households, rural households were more likely to implement each of the response strategies, even after adjusting for demographic, economic, and social characteristics as well as pandemic-related hardships.

The findings from this study are specific to these communities and are not generalizable to other contexts, but they are consistent with findings reported in other studies from Africa [5, 7, 8, 11, 12, 15, 20]. While we acknowledge this limitation, we note that it is common to studies using HDSS data [19]. As an observational study, other potential limitations include unmeasured variables (for example, household participation in Ethiopia’s Productive Safety Net Program (PSNP) and recall bias due to the extended length of time considered in the study (i.e., some respondents may have forgotten hardships that occurred closer to onset of the pandemic). As a cross-sectional study, we cannot draw causal inferences and we are likewise unable to explicitly model whether hardships experienced were due to COVID-19 or more generally due to pre-existing poverty and vulnerability, though we have endeavored to adjust for these factors in our models. Results from other studies suggest hardships associated with the COVID-19 pandemic are temporary [10]. Health and Demographic Surveillance Systems, like the one used here, offer the opportunity to collect longitudinal data to evaluate the impact of hardship events over time: follow-up data collection using the same survey instrument can easily be attached to subsequent rounds of data collection already being fielded. As is the case with other studies involving humanitarian crises, we are unable to differentiate between the effects of the pandemic and those of the political tension in the northern region of Ethiopia [30]. However, it is noteworthy that the northern region is more than 400 km away from the study communities.

## Conclusion

Households living in rural Eastern Ethiopia were at greater risk of experiencing a variety of hardships during the COVID-19 pandemic compared to urban households, even after adjusting for demographic, economic, and social risk factors. Moreover, the strength of the associations between household hardships and demographic, economic, and social risk factors was greater for rural households compared to urban ones. These results suggest that socioeconomic differences between urban and rural areas may have been amplified during the pandemic. Interventions to ameliorate the consequences of lockdowns and other efforts to stem disease spread should also consider place inequalities and differential vulnerabilities.

## Data Availability

All data produced in the present study are available upon reasonable request to the authors

## Appendix A

**Table A.1:**
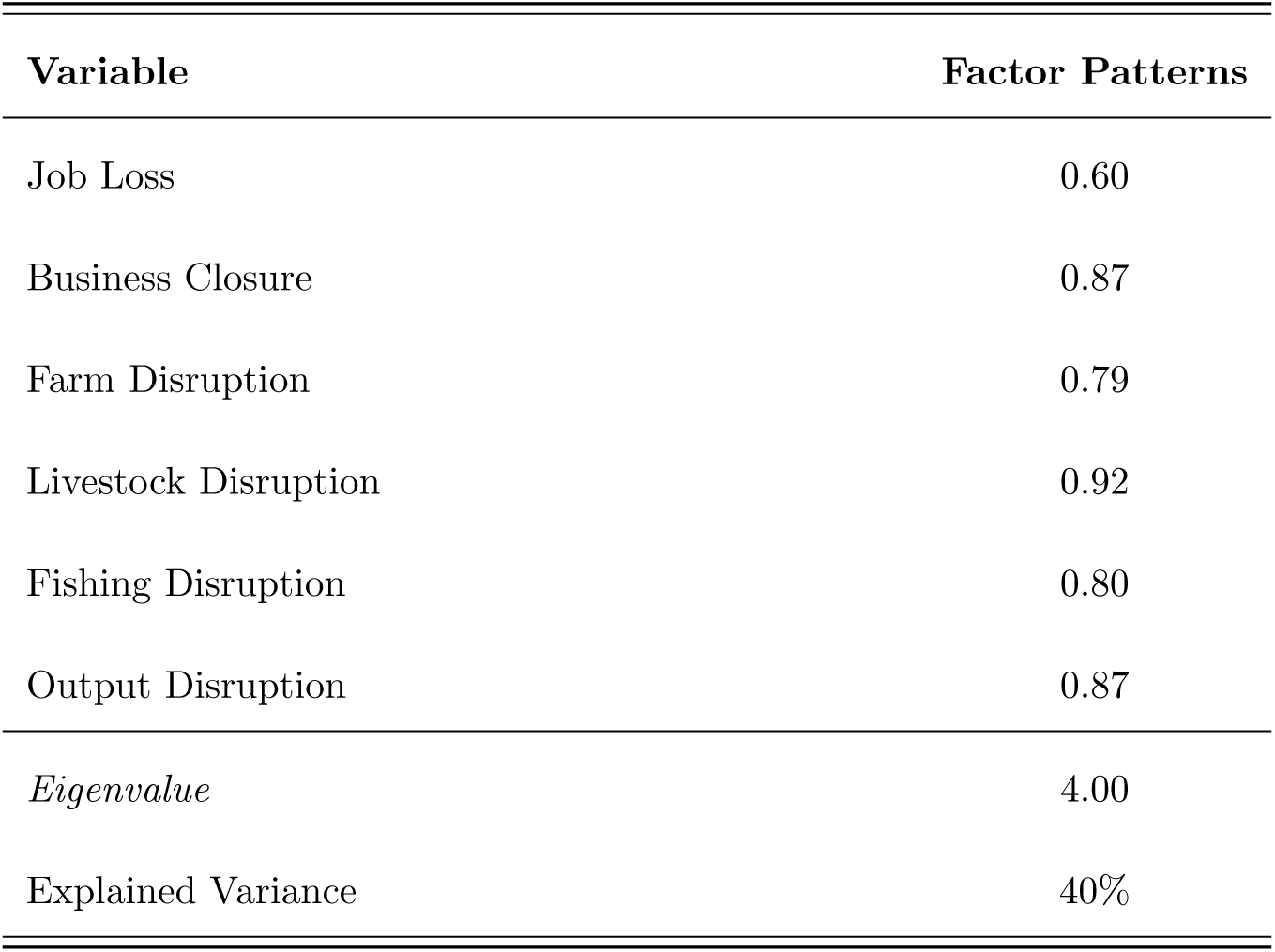
Principal Components Factor Analysis of Hardship Variables.

## Appendix B

A wealth index was generated based on a collection of assets and construction materials for the main dwelling of a given household. To generate the index, we followed recommendations from the DHS and the World Food Programme (WFP) that summarize steps for calculating an asset-based wealth index [48–50], including coding instructions for Stata [51], and an adaptation of this coding process implemented in R [30, 52], Using these documents to guide us, we generated our wealth index by identifying a list of household assets for inclusion in our index computation. We recoded all household assets into dichotomous variables and recoded dwelling materials into improved vs. non-improved dichotomous variables based on DHS recommendations. We divided our sample into rural and urban sub-samples and assessed the level of representation of a given variable within the rural and urban sub-samples (per WFP recommendations, a given variable was included in further calculations if percent ownership ranged between 5 and 95 percent). We then employed principal components analysis with varimax rotation to calculate component scores for those households living in either rural or urban areas, which explained 45% of the variation in both sub-samples, and extracted and combined the PCA scores of the first component from the urban and rural sub-samples; and finally organized the scores into wealth quintiles to generate a composite asset index. The distribution of assets owned by households as well as the materials used for constructing a household’s residence are presented in Figure B.1.

**Figure B.1.**
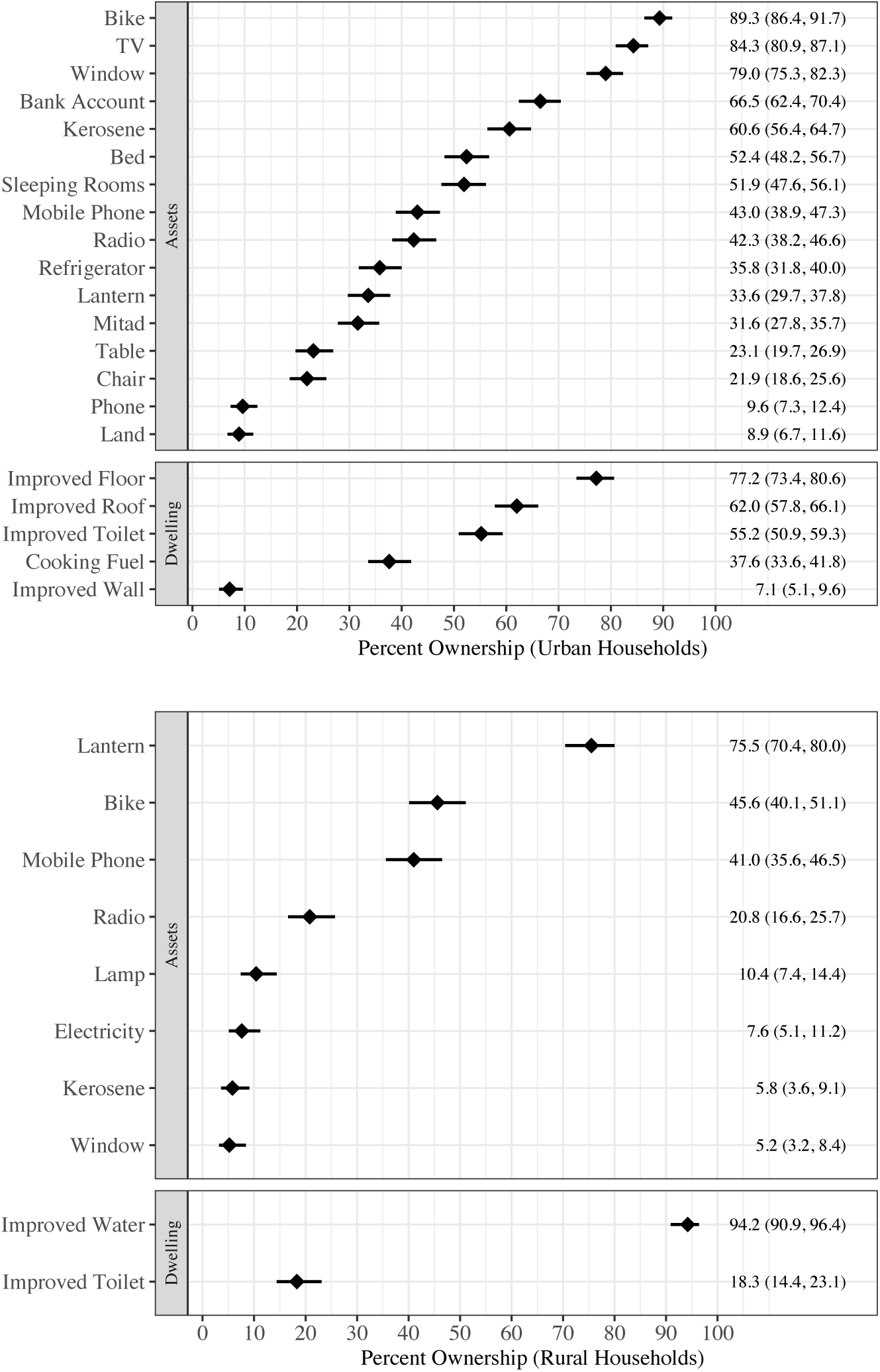
Distribution of Households’ Asset Ownership and Dwelling Construction Materials. These assets and construction materials were used as inputs for calculating the wealth index.

## Appendix C

**Figure C.1.**
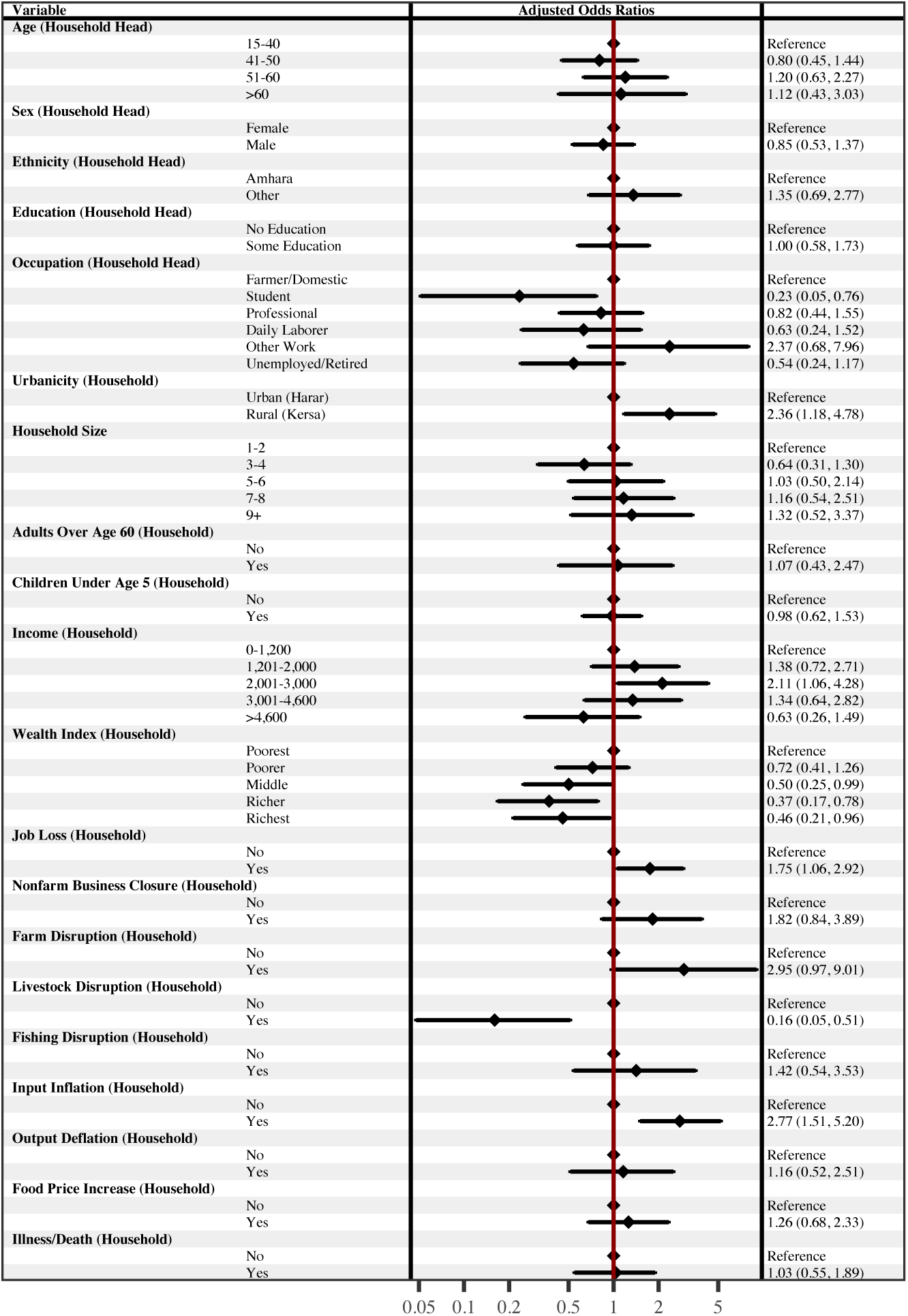
Adjusted Associations with Reduced Food Consumption presented as Adjusted Odds Ratios (AOR). The forest plot presents AORs with 95% confidence intervals from an adjusted logistic regression model. The AORs were adjusted for the other variables included in the model. Education and Occupation had 9 missing values; Income had one outlier set to missing. (n=756)

**Figure C.2.**
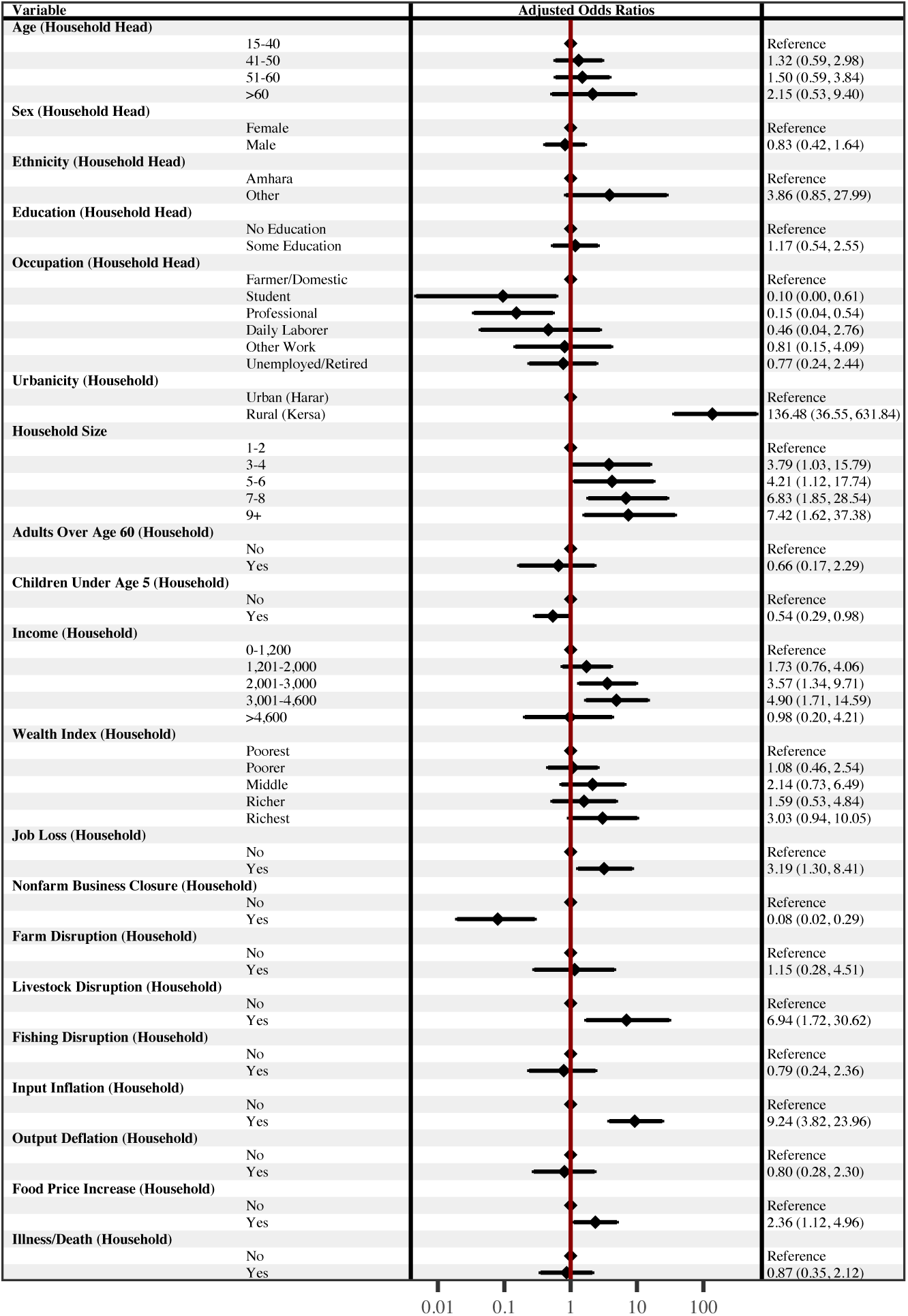
Adjusted Associations with Sold Assets presented as Adjusted Odds Ratios (AOR). The forest plot presents AORs with 95% confidence intervals from an adjusted logistic regression model. The AORs were adjusted for the other variables included in the model. Education and Occupation had 9 missing values; Income had one outlier set to missing. (n=756)

**Figure C.3.**
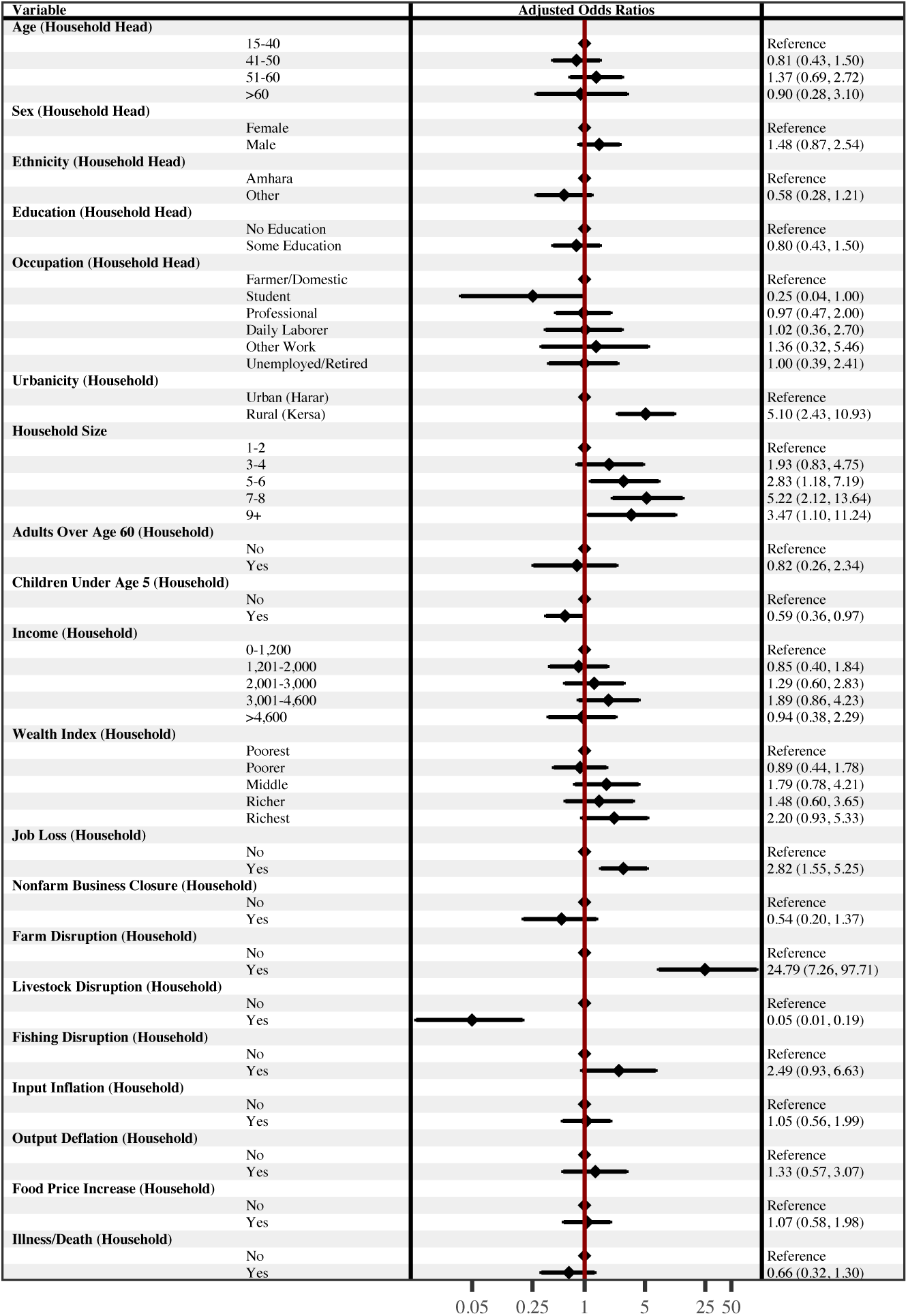
Adjusted Associations with Sought to Generate Additional Income presented as Adjusted Odds Ratios (AOR). The forest plot presents AORs with 95% confidence intervals from an adjusted logistic regression model. The AORs were adjusted for the other variables included in the model. Education and Occupation had 9 missing values; Income had one outlier set to missing. (n=756)

**Figure C.4.**
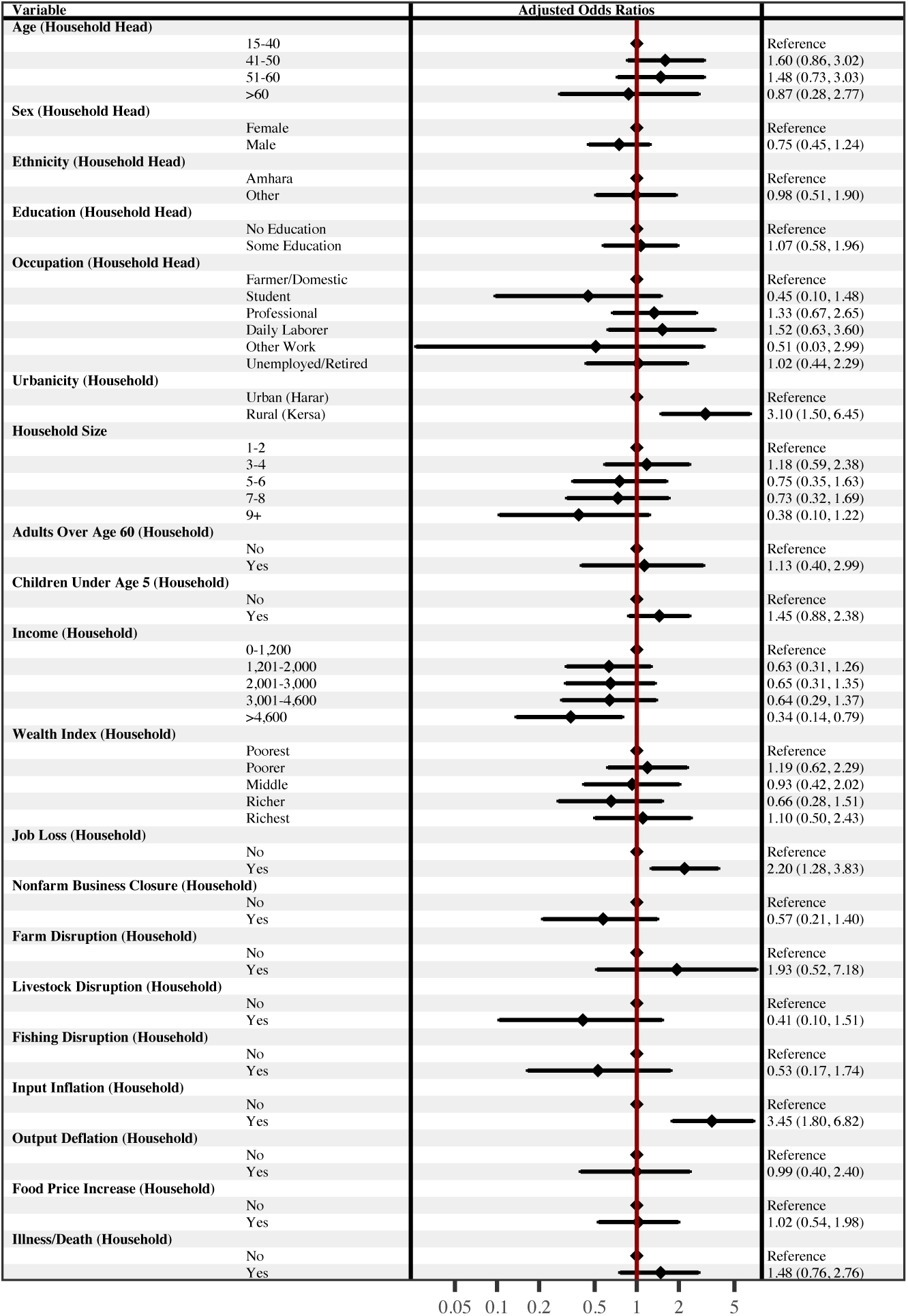
Adjusted Associations with Sought Help from Family or Friends presented as Adjusted Odds Ratios (AOR). The forest plot presents AORs with 95% confidence intervals from an adjusted logistic regression model. The AORs were adjusted for the other variables included in the model. Education and Occupation had 9 missing values; Income had one outlier set to missing. (n=756)

**Figure C.5.**
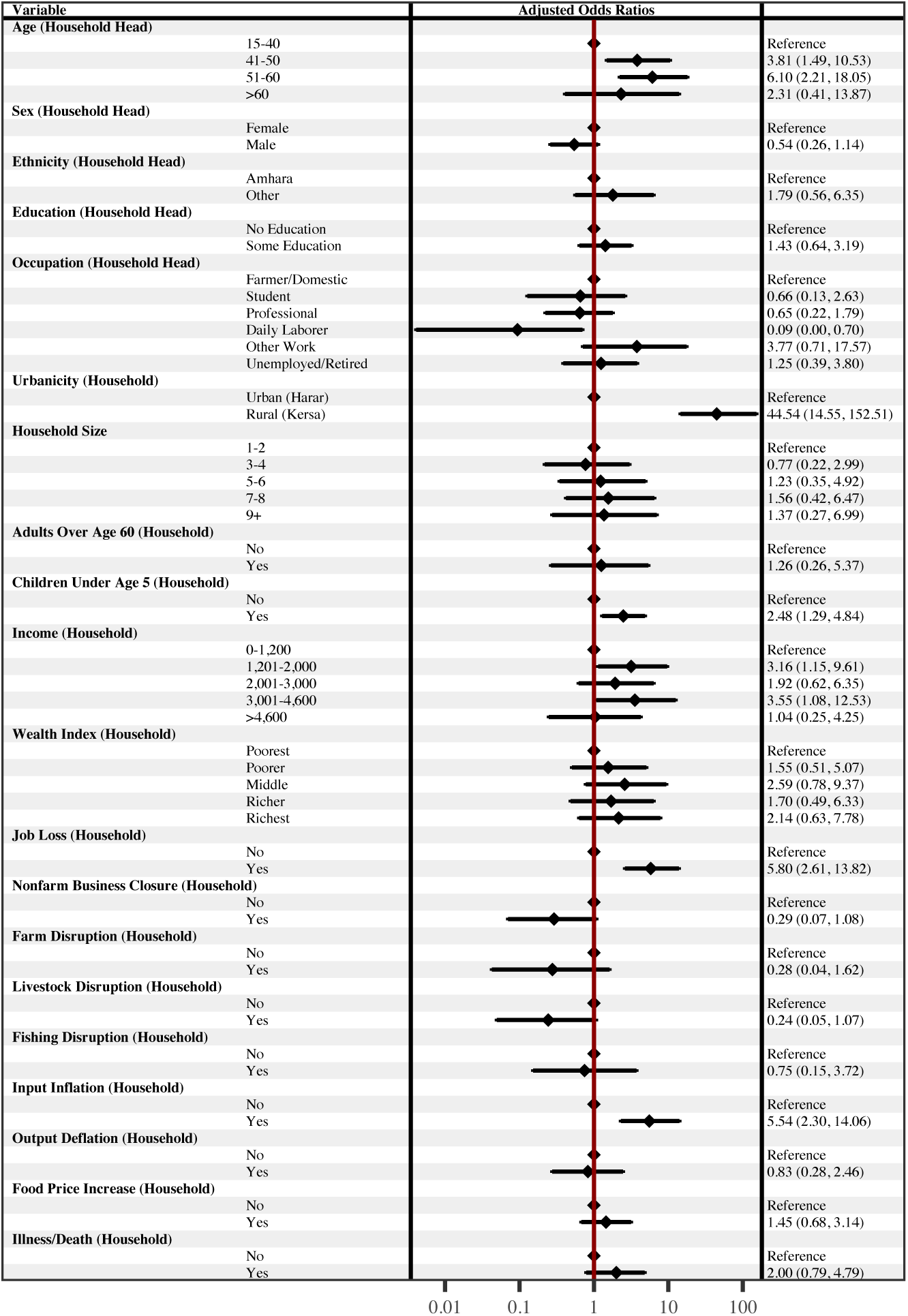
Adjusted Associations with Borrowed from Family or Friends presented as Adjusted Odds Ratios (AOR). The forest plot presents AORs with 95% confidence intervals from an adjusted logistic regression model. The AORs were adjusted for the other variables included in the model. Education and Occupation had 9 missing values; Income had one outlier set to missing. (n=756)

## Acknowledgements

We are grateful to the study participants who contributed their time in responding to our survey. We are also indebted to the fieldworkers in the data collection team that contacted household representatives and collected to data presented herein.

## Availability of data and materials

The data from the Ethiopia COVID-19 lockdown questionnaire are publicly available through the CHAMPS Population Surveillance Dataverse [39]. The HDSS data and code used for the analysis presented in this study are available upon reasonable request. HDSS data requests should be sent to.

## Ethics approval and consent to participate

This study was conducted according to the guidelines in the Declaration of Helsinki; all procedures involving research study participants, including digital data collection using tablets that were programmed with the corresponding survey instruments, were approved by the Institutional Health Research Ethics Review Committee (IHRERC), College of Health and Medical Sciences, Harar Campus, Ethiopia; approval reference number Ref.No.IHRERC/127/2021. Written informed consent was obtained for participants who were able to read and write. For participants who were unable to read or write, the informed consent statement was read and oral informed consent from the participant was obtained, recorded, and witnessed. These procedures for obtaining written or oral informed consent were approved by the Institutional Health Research Ethics Review Committee (IHRERC), College of Health and Medical Sciences, Harar Campus, Ethiopia; approval reference number Ref.No.IHRERC/127/2021.

## Consent for publication

NA

## Authors’ contributions

Conceptualization, J.M., S.C., N.A., and M.D.; methodology, J.M., S.C., N.A., and M.D.; software, J.M.; validation, J.M., Z.M., N.A., and M.D.; formal analysis, J.M. and Z.M.; investigation, J.M., Z.M., S.C., N.A., T.G., G.M., G.D., and M.D.; resources, J.M., Z.M., C.W., S.C., N.A., T.G., G.M., and M.D.; data curation, N.A., T.G., G.M., G.D., and M.D.; writing - original draft preparation, J.M.; writing - review and editing, J.M., Z.M., C.W., S.C., N.A., T.G., G.M., G.D., and M.D.; visualization, J.M. and Z.M.; supervision, C.W., S.C., and N.A.; project administration, J.M. and M.D.; funding acquisition, C.W.

